# Mortality rate and estimate of fraction of undiagnosed COVID-19 cases in the US in March and April 2020

**DOI:** 10.1101/2020.05.28.20116095

**Authors:** Ru-Shan Gao, Karen Rosenlof

## Abstract

We use a simple model to derive a mortality probability distribution for a patient as a function of days since diagnosis (considering diagnoses made between 25 February and 29 March 2020). The peak of the mortality probability is the 13^th^ day after diagnosis. The overall shape and peak location of this probability curve are similar to the onset-to-death probability distribution in a case study using Chinese data.

The total mortality probability of a COVID-19 patient in the US diagnosed between 25 February and 29 March is about 21%. We speculate that this high value is caused by severe under-testing of the population to identify all COVID-19 patients. With this probability, and an assumption that the true probability is 2.4%, we estimate that 89% of all SARS-CoV-2 infection cases were not diagnosed during this period.

When the same method is applied to data extended to 25 April, we found that the total mortality probability of a patient diagnosed in the US after 1 April is about 6.4%, significantly lower than for the earlier period. We attribute this drop to increasingly available tests. Given the assumption that the true mortality probability is 2.4%, we estimate that 63% of all SARS-CoV-2 infection cases were not diagnosed during this period (1 – 25 April).

## 1. Introduction

Severe acute respiratory syndrome coronavirus 2 (SARS-CoV-2), as of 27 April 2020, has sickened at least 3 million people including, in the US alone, over 1 million diagnosed COVID-19 cases. Lacking either a vaccine or effective treatments, containing and combating the disease requires mass-testing potential patients followed by quarantine and social distancing. It is widely believed that the lack of testing capability exacerbated the COVID-19 pandemic in the USA. A reliable estimate of the severity of the missed diagnosis problem cannot be made now even with massive testing of antibodies to COVID-19.

Here we use publicly available data of daily COVID-19 cases and deaths in the US (e.g. https://www.worldometers.info/coronavirus/country/us/) to calculate the mortality rate of diagnosed patients in the USA. Comparing this rate with known low mortality rates of other countries with better testing rates leads to an estimate of fractional undiagnosed cases.

## 2. Methods

### 2.1 Calculating mortalities from daily diagnosis

On a given day *x* a certain number of new cases NC(*x*) are diagnosed. Each diagnosed patient has a certain probability P(*d*) of dying every day after his/her diagnosis. The mortality rate m(*d*) of patients on day *d* is

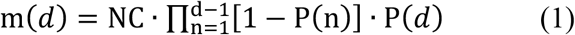

Where n = 1 denotes the starting day of dataset, and the P factor represents fraction of patients still alive on this day. The mortality rate M(*k*) of all surviving patients on any day *k* is then

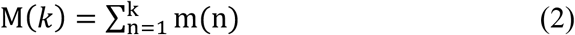

P(*d*) is unknown. We use reported daily NC_r_(d) and M_r_(d) (https://www.worldometers.info/coronavirus/country/us/) to derive P(y), the probability of dying on a given day of infection. Here subscript r denotes real data. We assume P(*d*) is a Gaussian for simplicity: P(*d*) = A·exp((*d-d_m_*)^2/w), where A (maximum probability), *d_m_* (date for the maximum probability), and w (width of the Gaussian peak) are Gaussian parameters and assumed to be constant throughout the study period. Using equations 1 and 2 A, *d_m_*, and w can be determined by fitting M(*k*) to M_r_.

### 2.2 Estimating the true COVID-19 mortality rate

The true COVID-19 mortality rate is needed to estimate the fraction of SARS-CoV-2 infection cases not diagnosed. The determination of the true COVID-19 mortality rate cannot be done without first diagnosing all COVID-19 cases, which has not occurred anywhere. We opt to estimate the upper limit of the mortality rate instead. This upper limit is defined as the ratio of total deaths to total diagnosed cases. Only countries/groups where almost all COVID-19 cases closed (> 90%) are used in our analysis. This is because total mortalities will continue to increase if a large fraction of cases are still active. A total of four countries/group meet this criterion: China, Diamond Princess Cruise Ship, Iceland, and Thailand. The upper limit of the mortality rates from these four are 0.056, 0.018, 0.0056, 0.018, respective. We take the average of these four values (0.024, or 2.4%) as an estimate of the true COVID-19 mortality rate. Note that this value is very close to the value of 2.3% by Wu et al. (2020).

## 3. Results

Fitting M(k) to M_r_(d) can be done rigorously using the least-square fitting (e.g. Gao et al., 1989). However, here we use manual optimization for simplicity.

### 3.1 Before March 29

We examine early data (25 February to 28 March) first, when the SARSCoV-2 infection tests were very limited. In the next section the analysis is extended to 14 April, with increased test availability.

Results based on early data are shown in Figures 1 and 2.

**Figure 1.**
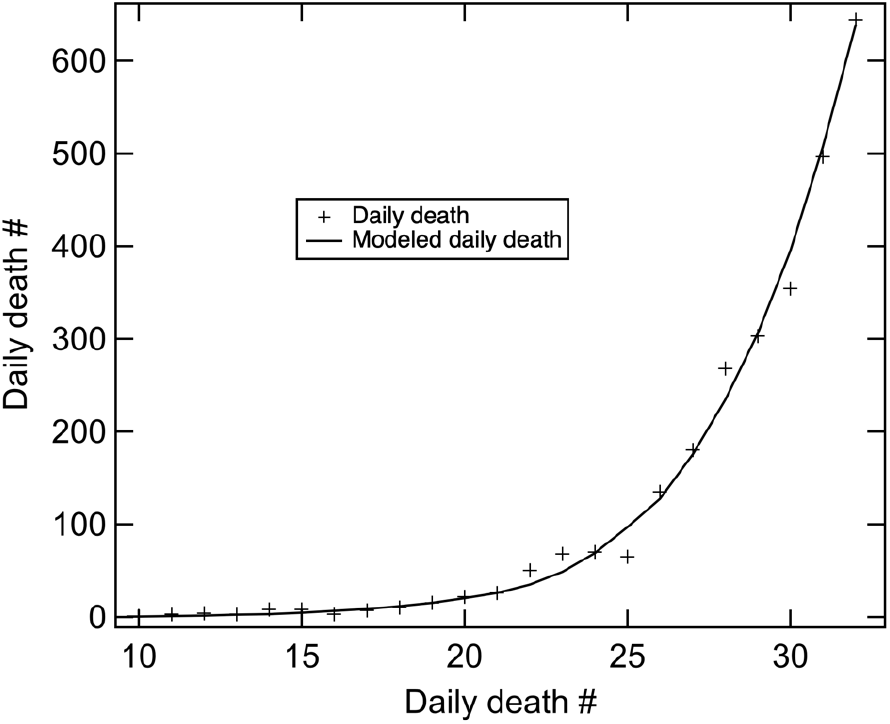
Real and modeled mortality (using Equations 1 and 2) rates. The fit mortality probability is shown in Figure 2.

**Figure 2.**
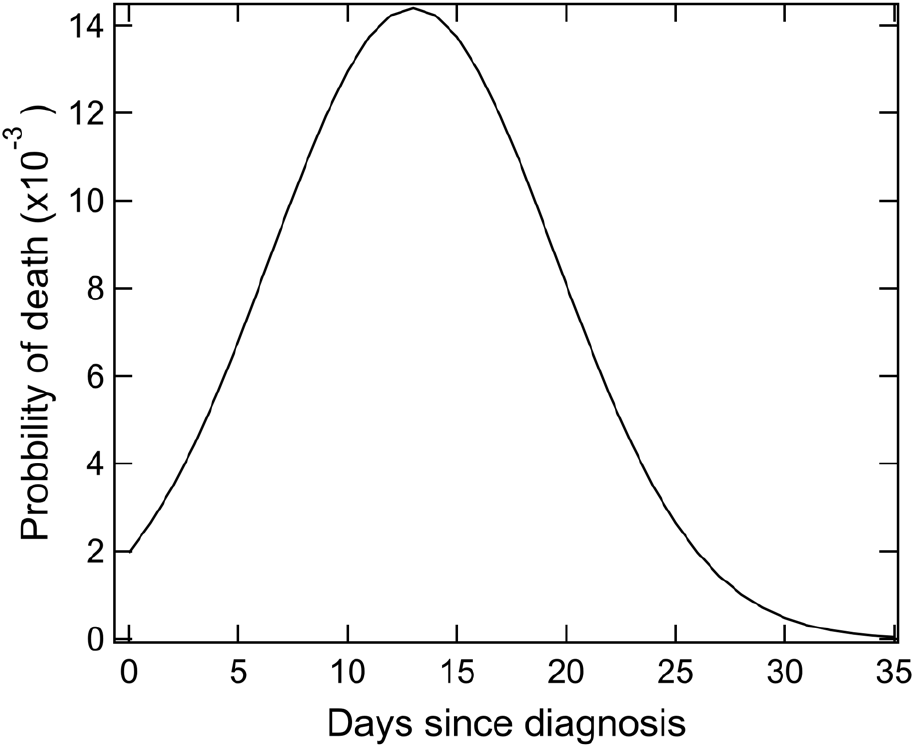
The fit mortality probability for a patient as a function of days since diagnosis (between 25 February and 29 March 2020).

The total mortality probability for a patient throughout the course of their COVID-19 encounter is 1 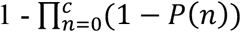, where P(*n*) is the mortality probability n days after diagnosis and c denote the course length in days. For the probability curve shown in Figure 2, the total mortality probability is 0.21, meaning patients diagnosed in this period (25 Feb – 28 Mar 2020) have a mortality rate of 21%. This is a very high number compared to the estimate of the true mortality rate (see Section 2.2). One likely cause of this high rate is that a high number of SARS-CoV-2 infection cases were not diagnosed. Assume that the true mortality rate in the USA is 2.4%, the 21% number in the USA suggests that 89% of all SARS-CoV-2 infection cases were not diagnosed during this period.

We note that the overall shape and peak location of probability curve shown in Figure 2 are similar to the onset-to-death probability distribution in a case study using Chinese data (Verity et al., 2020).

### 3.2 Extending to 25 April

When the same calculation is extended to a later day (25 April), the model significantly overpredicts daily mortality rate (Figure 3, black curve). This overprediction is understandable since the availability of SARS-CoV-2 tests increased, thereby allowing a larger fraction of cases to be diagnosed. Therefore, using the same mortality probability function derived from earlier data is not appropriate. To address this problem, we introduce a mortality probability distribution modifier (Figure 4). By adjusting the shape of this function, we are able to match the reported daily mortalities again (red curve in Figure 3). Figure 5 shows the total mortality probability for each patient as function of date when the case had been diagnosed. As shown, the total probability decreases dramatically from an initial value of 21% to 6.4%. Again, using an assumed true mortality rate of 2.4%, the data shown in Figure 5 suggests that about 63% of all SARS-CoV-2 infection cases were not diagnosed in the period between 1 and 25 April.

**Figure 3.**
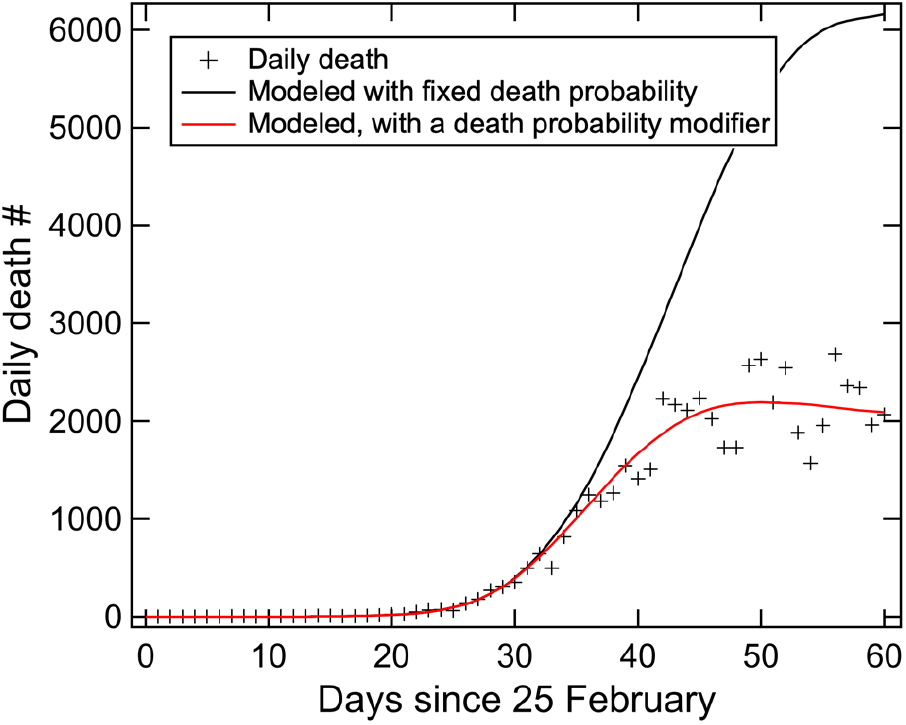
Real (crosses) and modeled mortality (black and red curves) rates. The black curve is derived using the mortality probability distribution shown in Figure 2. The red curve is derived using a mortality probability distribution modifier function shown in Figure 4.

**Figure 4.**
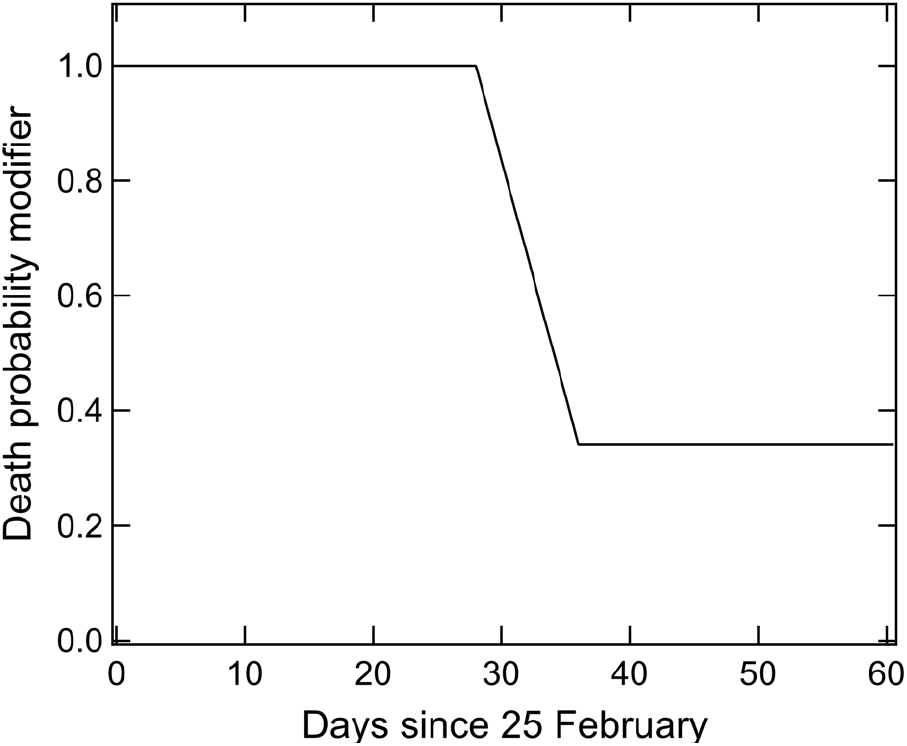
Mortality probability distribution modifier (multiplicative) is shown as a function of diagnosis date.

**Figure 5.**
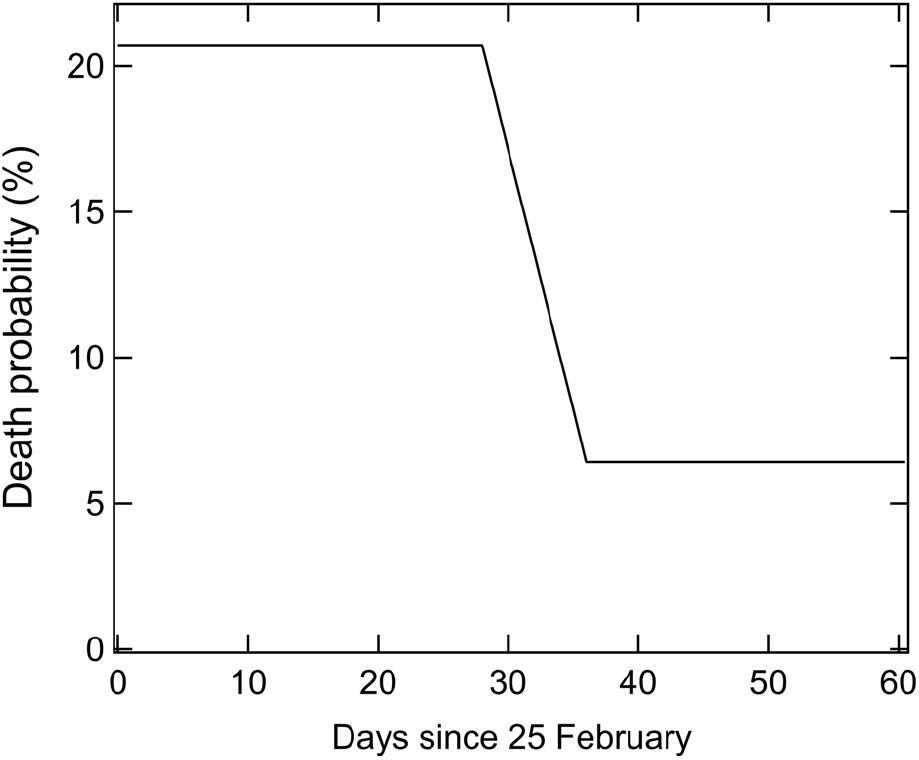
Total mortality probability with the modifier shown in Figure 4 as a function of diagnosis date.

## 4. Discussion

The total US mortality probability after 1 April is about 6.4%. This value is slightly higher than the apparent mortality probability (total mortalities/total cases) as of 25 April (5.6%). These two values are not inconsistent with each other, since the apparent mortality probability is necessarily lower than the real mortality probability. This is because the highest mortality probability occurs two weeks after diagnosis (Figure 2). Total deaths would still climb even if there were no new cases after 25 April.

We note that the probability modifier shown in Figure 4 is an oversimplification of the real situation. However, the relatively sharp transition between 25 March and 1 April (Day 29 – 36) is critical to achieve a fit of the mortality data (red curve in Figure 3). This transition suggests that tests became more readily available during this period.

For simplicity we lumped the data from the entire USA for our analysis. In doing so, some state-by-state details are lost. It would be interesting to extend our analysis to each state, and to different countries. Due to the lack of effective treatments, the overall shape of the mortality probability curve shown in Figure 2 is likely uniform throughout the country although the absolute values will vary state-by-state.

## Data Availability

The data analyzed came from the website www.worldometers.info, and this is publicly available.

https://www.worldometers.info/coronavirus/country/us/

## Acknowledgments

We thank David W. Fahey for insightful comments on an earlier version of this manuscript.

